# Improved characterization of gene-environment interactions for vitamin D through variance quantitative trait loci

**DOI:** 10.1101/2024.08.30.24312868

**Authors:** Tianyuan Lu, Wenmin Zhang, Cassianne Robinson-Cohen, Corinne D. Engelman, Qiongshi Lu, Ian H. de Boer, Lei Sun, Andrew D. Paterson

**Affiliations:** Department of Population Health Sciences, University of Wisconsin-Madison, Madison, WI, USA; Department of Biostatistics and Medical Informatics, University of Wisconsin-Madison, Madison, WI, USA; Department of Statistical Sciences, University of Toronto, Toronto, ON, Canada; Montreal Heart Institute, Montreal, QC, Canada; Division of Nephrology and Hypertension, Department of Medicine, Vanderbilt University Medical Center, Nashville, TN, USA; Department of Statistics, University of Wisconsin–Madison, Madison, WI, USA; Center for Demography of Health and Aging, University of Wisconsin–Madison, Madison, WI, USA; Division of Nephrology, Department of Medicine, University of Washington, Seattle, WA, USA; Kidney Research Institute, University of Washington, Seattle, WA, USA; Division of Biostatistics, Dalla Lana School of Public Health, University of Toronto, Toronto, ON, Canada; Division of Epidemiology, Dalla Lana School of Public Health, University of Toronto, Toronto, ON, Canada; Genetics and Genome Biology, The Hospital for Sick Children, Toronto, ON, Canada

## Abstract

**Background:** Understanding gene-environment interaction effects influencing vitamin D status may refine nutrition and public health strategies for vitamin D deficiency. Recent methodological advances have enabled the identification of variance quantitative trait loci (vQTLs) where gene-environment interaction effects are enriched.

**Objectives:** To identify vQTLs for serum 25-hydroxy vitamin D (25OHD) concentration and characterize potential gene-environment interaction effects of vQTLs.

**Methods:** We conducted vQTL discovery for 25OHD using a newly developed quantile integral linear model in the UK Biobank individuals of European (N = 313,514), African (N = 7,800), East Asian (N = 2,146), and South Asian (N = 8,771) ancestries, respectively. We tested for interaction effects between the identified vQTL lead variants and 18 environmental, biological, or lifestyle factors, followed by multiple sensitivity analyses.

**Results:** We identified 19 independent vQTL lead variants (p-value <5×10^-8^) in the European ancestry population. No vQTLs were identified in the non-European ancestry populations, likely due to limited sample sizes. A total of 32 interaction effects were detected with a false discovery rate <0.05. While known gene-season of measurement interaction effects were confirmed, additional interaction effects were identified involving modifiable risk factors, including time spent outdoors and body mass index. The magnitudes of these interaction effects were consistent within each locus upon adjusting for season of measurement and other covariates. We also identified a gene-sex interaction at a vQTL that implicates *DHCR7*. Integrating transcript- and protein-level evidence, we found that the sex-differentiated genetic effects may act through sex-biased expression of DHCR7 isoforms in skin tissues due to alternative splicing.

**Conclusions:** Through the lens of vQTLs, we identified additional gene-environment interaction effects affecting vitamin D status apart from season of measurement. These findings may provide new insights into the etiology of vitamin D deficiency and encourage personalized prevention and management of associated diseases for at-risk individuals.

**Supplementary Tables are available at https://figshare.com/s/82f05d4c830dc4af50ac**

## Background

Vitamin D is an essential vitamin that has important roles in the musculoskeletal, nervous, and immune systems^1,2^. Vitamin D has multiple forms, such as cholecalciferol (D_3_), ergocalciferol (D_2_), 25-hydroxy vitamin D (25OHD), and calcitriol. A sufficient amount of vitamin D, often measured using serum concentrations of 25OHD^3^, is required for maintaining calcium and phosphorus homeostasis, regulating cellular proliferation and differentiation, and modulating immune functions. Long-term vitamin D deficiency can lead to rickets in children and osteomalacia in adults^4^, and has been associated with various cardiometabolic, neurocognitive, and autoimmune diseases^5^.

Vitamin D in the form of cholecalciferol can be synthesized endogenously when the skin is exposed to ultraviolet rays, which act on 7-dehydrocholesterol^2,3^. Vitamin D can also be obtained in the forms of cholecalciferol or ergocalciferol from dietary intake, such as oily fish, fortified foods, and vitamin D supplementation^6^. Both cholecalciferol and ergocalciferol are hydroxylated in the liver to be converted into 25OHD^2,3^. 25OHD is further hydroxylated, primarily in the kidneys, to produce calcitriol, the biologically active form of vitamin D^2,3^. Large-scale genome-wide association studies (GWASs) have consistently identified genetic variants associated with serum 25OHD concentrations that map to genes with known roles in vitamin D metabolism^7–10^. These canonical vitamin D genes include *GC* (vitamin D-binding protein), *CYP2R1* (catalyzing the hydroxylation of cholecalciferol and ergocalciferol), *DHCR7* (7-dehydrocholesterol reductase), and *CYP24A1* (catalyzing the catabolism of calcitriol).

Since vitamin D status is influence by both genetic and environmental factors, characterizing potential gene-environment interaction effects is crucial for better understanding the etiology of vitamin D deficiency (often defined as a serum 25OHD concentration below 20 ng/mL or 50 nmol/L^3^) and exploring more effective approaches for prevention and early intervention. Existing studies have shown that indicators of sunlight exposure, such as season of measurement, have strong interaction effects with several genetic variants on serum 25OHD concentrations^7,8,11–13^, such as those mapping to *GC* and *CYP2R1*. However, it remains challenging to characterize potential interaction effects between genetic variants and other risk factors since genome-wide interaction analyses entail a high multiple testing burden, while the interaction effects often account for only a small proportion of the heritability of polygenic traits^14–16^.

An alternative approach is to identify variance quantitative trait loci (vQTLs), where the genotypes are associated with the variance heterogeneity of a trait, without necessarily being associated with the mean^14,15^. It has been shown that gene-environment interaction effects are enriched in vQTLs because unmodeled interaction effects contribute to the genotype-associated variance heterogeneity^14,15^. Therefore, tests for gene-environment interaction effects can focus on vQTLs rather than the whole genome to reduce the multiple testing burden. Several methods have been developed and used for vQTL identification^14,17–19^. For instance, a recent study applied the median-based Levene’s test to identify vQTLs for serum 25OHD concentrations and replicated known gene-season of measurement interaction effects^7^. A newly developed quantile integral linear model (QUAIL) has demonstrated substantially improved statistical power in vQTL identification over previous methods^17^, but its utility has not been extensively explored.

In this work, we applied QUAIL to identify vQTLs for serum 25OHD concentrations in the UK Biobank^20^ (N = 313,514 European ancestry, 7,800 African ancestry, 2,146 East Asian ancestry, and 8,771 South Asian ancestry individuals). We tested for interaction effects between vQTL lead variants and 18 environmental, biological, or lifestyle factors. We conducted multiple sensitivity analyses to examine the robustness of our results and integrated transcript- and protein-level evidence to investigate potential biological mechanisms. Our findings improve the understanding of gene-environment interaction effects affecting vitamin D status and may provide novel insights into personalized health interventions.

## Methods

### UK Biobank

From 2006 to 2010, the UK Biobank recruited approximately 500,000 individuals at 22 assessment centers in the United Kingdom^20^. Upon recruitment, medical history, environmental factors, and lifestyle characteristics were collected by interviews or questionnaires. The participants underwent multiple physical measurements and provided biological samples for further investigations. The sample handling and storage protocol has been described previously^20^. Serum 25OHD concentrations were measured using Chemiluminescence Immunoassay (DiaSorin Liaison XL) with an analytical range of 10-375 nmol/L. The quality of serum 25OHD measurements was externally verified following the RIQAS Immunoassay Specialty 1 scheme.

Genome-wide genotyping was performed using the Applied Biosystems UK BiLEVE Axiom Array or UK Biobank Axiom Array. The genotypes were imputed using the Haplotype Reference Consortium and the merged UK10K and 1000 Genomes phase 3 reference panels^21,22^. We retained genetic variants with a minor allele frequency >5% and an imputation quality score >0.8. Genetic ancestry assignment was performed based on the clustering of genetic principal components and mapping to the 1000 Genomes Project super-populations as described previously^23,24^. Within each genetic ancestry, pairwise genetic relatedness was estimated using GCTA^25^ (version 1.93.3beta2). We randomly excluded one individual from each pair of individuals with a coefficient of relationship ≥0.125, which corresponds to a third-degree or closer relationship.

### Discovery of variance quantitative trait loci

We conducted ancestry-specific genome-wide vQTL analyses for serum 25OHD using QUAIL^17^. For a quantitative trait, QUAIL derives quantile integrated rank scores and performs linear regression with the quantile integrated rank scores as the outcome within a GWAS framework to identify vQTLs^17^. Raw serum 25OHD concentrations were first converted into quantile integrated rank scores. Following practical recommendations^17^, we used the default 2,000 quantile levels for the European ancestry population and 500 quantile levels for the non-European ancestry populations with smaller sample sizes. Age, age^2^, sex, age x sex, age^2^ x sex, genotyping array, assessment center, season of measurement, and the first 10 genetic principal components (separately generated in each population) were included as covariates. The season of measurement was determined by the month of blood sample collection^8^: winter (January-March), spring (April-June), summer (July-September), and fall (October-December). Since the dose of vitamin D supplementation was unavailable in the UK Biobank data, individuals who self-reported being on vitamin D supplementation (data fields 6155 and 20084) were included in sensitivity analyses but not in primary analyses.

For autosomal variants, GWAS of the quantile integrated rank scores was performed while adjusting for the same covariates. For variants on chromosome X, we conducted sex-stratified GWAS, adjusting for all covariates except sex, age x sex, and age^2^ x sex, followed by sample size-based meta-analysis to assess statistical significance^26^. We also conducted a GWAS of the raw serum 25OHD concentrations to identify quantitative trait loci (QTLs) that are significantly associated with the mean but not necessarily with the variance of the outcome. Quantile-quantile (Q-Q) plots were used to examine whether population stratification and other confounding factors were properly controlled.

Since no vQTLs were identified in the non-European ancestry populations, further analyses were restricted to the European ancestry population. We performed linkage disequilibrium (LD) clumping to identify independent lead variants of vQTLs with a p-value <5.0×10^-8^ and an LD r^2^ <0.001. We further performed colocalization analyses using SharePro^27^ with a default prior colocalization probability of 1.0×10^-5^ to evaluate whether the same genetic variants influenced both the mean and variance of serum 25OHD concentrations. QTL and vQTL summary statistics for variants located within 1Mb of each lead variant were used to assess colocalization, where overlapping ±1Mb regions were merged into one locus. For LD clumping and colocalization analyses, the LD structures were obtained based on all European ancestry individuals included in GWAS.

### Functional annotation of variance quantitative trait loci

For each vQTL lead variant, we queried the Open Targets Genetics database^28^ (version 22.10) to obtain its functional annotations, including predicted functional impacts on nearby genes and annotations of expression, splicing, and circulating protein quantitative trait loci (eQTLs, sQTLs, and pQTLs, respectively). We also retrieved phenome-wide associations (p-value <5.0×10^-8^) with the vQTL lead variants reported in the NHGRI/EBI GWAS Catalog, UK Biobank, or FinnGen from the Open Targets Genetics database^28^ (version 22.10).

### Sensitivity analyses

We performed multiple sensitivity analyses to evaluate the robustness of our results. First, for each vQTL lead variant, we performed conditional quantile regression with the raw serum 25OHD concentrations and inverse rank normal transformed serum 25OHD concentrations, respectively, at a series of quantiles (0.05, 0.10, …, 0.90, 0.95), and examined whether the trend of varying genetic effects was consistent. Second, for each vQTL lead variant, we applied alternative methods for homogeneity of variance test, including median-based Levene’s test, Fligner-Killeen’s test, and location and scale tests^29,30^, after regressing out the effects of covariates included in GWAS on serum 25OHD concentrations. The location and scale tests assess both additive and dominance effects on the mean and variance, respectively. An F-test in the scale test stage is equivalent to a mean-based Levene’s test^29^. Third, we included European ancestry individuals who self-reported being on vitamin D supplementation. For these individuals, we subtracted 21.2 nmol/L from their measured serum 25OHD concentrations, which corresponds to the average effect of taking 400 international units of cholecalciferol per day^8^. We then repeated genome-wide vQTL discovery using QUAIL.

### Detection of gene-environment interaction effects

For each vQTL lead variant, we tested for potential interaction with measured environmental factors with raw serum 25OHD concentrations as the outcome. The candidate environmental factors included season of measurement (defined above), age, sex, body mass index (BMI; data field 21001), waist-hip ratio (WHR; derived using data fields 48 and 49), smoking status (data field 20116), alcohol intake, time spent outdoors (average of data fields 1050 and 1060), food intake, estimated glomerular filtration rate (eGFR; derived using the Chronic Kidney Disease Epidemiology Collaboration 2021 Formula^31^ with serum creatinine; data field 30700), serum alanine transaminase concentrations (ALT; data field 30620), serum aspartate transaminase concentrations (AST; data field 30650), and serum gamma glutamyl transferase concentrations (GGT; data field 30730).

Alcohol intake was quantified by estimating the weekly intake of pure alcohol^32,33^ (1 pint of beer or cider = 6 grams, 1 standard glass of white or red wine = 16.8 grams, 1 glass of fortified wine = 14.08 grams, 1 shot of spirits or liquor = 8 grams, and 1 glass of other alcohol drinks = 12 grams). This was then converted into standard drinks (1 unit = 14 grams of pure alcohol).

Food intake was quantified by 17 dietary components (category 100052). We performed principal component analysis of these food intake variables after median imputation and standardizing each variable to have a unit variance. We included principal components that captured more than 1/17 of the total variance as candidate environmental factors.

For each pair of lead variant and candidate environmental factor, we fitted a baseline model that included the main effects of the lead variant and the environmental factor, and a full model that included these main effects along with the variant-environmental factor interaction effect. Covariates (if not already a candidate environmental factor) in both models included age, age^2^, sex, age x sex, age^2^ x sex, genotyping array, assessment center, season of measurement, and the first 10 genetic principal components. The significance of interaction effects was assessed using a likelihood ratio test comparing the full model to the baseline model. Sensitivity analyses were conducted with inverse rank-normal transformed serum 25OHD concentrations as the outcome. Interaction effects with a Benjamini-Hochberg-corrected p-value (false discovery rate, FDR) <0.05 were considered significant while those with a Bonferroni-corrected p-value <0.05 were prioritized for interpretation.

Furthermore, for each of the lead variants that demonstrated significant interaction effects with two or more candidate environmental factors, we fitted a multiple-interactions model including the main effects of the lead variant and all involved environmental factors, all variant-environmental factor interaction effects, and the same covariates. We evaluated whether the estimated interaction effects were consistent upon the inclusion of other environmental factors.

### Investigation of sex-differentiated genetic effects

We aggregated additional lines of evidence to better understand the variant-sex interaction effect detected exclusively at rs732934. We obtained tissue-specific eQTL and sQTL summary statistics and results of sex-differential gene expression analyses from the Genotype-Tissue Expression (GTEx) project^34^. Details of the GTEx resources and analyses have been described previously^34–36^. In each tissue, genes with a local false sign rate (LFSR) <0.05 inferred using multivariate adaptive shrinkage were considered to have sex-differentiated expression^35^. For protein isoforms resulting from alternative splicing, we used InterPro^37^ (version 90.0) to predict their structural and functional domains using the amino acid sequences.

## Results

### Serum 25OHD variance quantitative trait loci

In this study, ancestry-specific vQTL analyses for serum 25OHD concentrations were based on 313,514 European ancestry, 7,800 African ancestry, 2,146 East Asian ancestry, and 8,771 South Asian ancestry participants, respectively. Demographic characteristics are summarized in **Table 1** and **Supplementary Table S1**.

**Table 1.** Characteristics of the UK Biobank European ancestry study population.

| *Characteristic* | *Participants (N = 313,514)* | *Missing data (%)* |
| --- | --- | --- |
| **Age (years)**, mean (SD) | 56.8 (8.0) | 0 |
| Median (IQR) | 58.0 (50.0-63.0) |  |
| **Sex**, No. (%) |  | 0 |
| Female | 164,158 (52.4%) |  |
| Male | 149,356 (47.6%) |  |
| **Serum 25OHD concentration (nmol/L)**, mean (SD) | 49.4 (20.9) | 0 |
| Median (IQR) | 47.7 (33.4-63.0) |  |
| **Season of measurement**, No. (%) |  | 0 |
| Fall | 70,473 (22.5%) |  |
| Winter | 76,833 (24.5%) |  |
| Spring | 91,578 (29.2%) |  |
| Summer | 74,630 (23.8%) |  |
| **Body mass index (kg/m<sup>2</sup>)**, mean (SD) | 27.4 (4.8) | 1,019 (0.3%) |
| Median (IQR) | 26.8 (24.2-29.9) |  |
| **Waist-hip ratio**, mean (SD) | 0.87 (0.09) | 545 (0.2%) |
| Median (IQR) | 0.88 (0.80-0.94) |  |
| **Smoking status**, No. (%) |  | 1,152 (0.4%) |
| Current smoker | 31,575 (10.1%) |  |
| Previous smoker | 110,163 (35.3%) |  |
| Never smoker | 170,624 (54.6%) |  |
| **Alcohol intake frequency (unit/week)**, mean (SD) | 9.2 (10.6) | 285 (0.1%) |
| Median (IQR) | 6.8 (0.8-13.5) |  |
| **Average time spent outdoors (hour)**, mean (SD) | 2.8 (2.0) | 20,385 (6.5%) |
| Median (IQR) | 2.5 (1.5-4.0) |  |
| **eGFR (mL/min/1.73m<sup>2</sup>)**, mean (SD) | 94.5 (13.0) | 383 (0.1%) |
| Median (IQR) | 97.1 (87.0-103.5) |  |
| **Serum ALT concentration (U/L)**, mean (SD) | 23.5 (14.0) | 303 (0.1%) |
| Median (IQR) | 20.2 (15.4-27.4) |  |
| **Serum AST concentration (U/L)**, mean (SD) | 26.2 (10.5) | 1,291 (0.4%) |
| Median (IQR) | 24.3 (21.0-28.8) |  |
| **Serum GGT concentration (U/L)**, mean (SD) | 37.0 (40.5) | 314 (0.1%) |
| Median (IQR) | 26.2 (18.5-40.7) |  |

25OHD: 25-hydroxy vitamin D
eGFR: Estimated glomerular filtration rate
ALT: Alanine transaminase
AST: Aspartate transaminase
GGT: Gamma glutamyl transferase

In the European ancestry population, consistent with previous studies^7,8^, GWAS of serum 25OHD concentrations revealed a polygenic architecture (**Figure 1A**). GWAS of the serum 25OHD quantile integrated rank scores identified 15 vQTLs, containing a total of 19 independent lead variants (**Figure 1B** and **Supplementary Table S2**). As expected, the mean effects (i.e. QTLs) and the variance effects (i.e. vQTLs) were correlated (**Supplementary Figure S1**). Colocalization analyses confirmed that the same genetic variants influenced both the mean and variance of serum 25OHD concentrations for 13 of the 15 vQTLs (posterior colocalization probability >0.99), including those implicating canonical vitamin D genes: *GC*, *CYP2R1*, *DHCR7*, and *CYP24A1* (**Figure 1C**). Notably, colocalization evidence was weak for the remaining two vQTLs where the mean effects were not genome-wide significant (**Figure 1C**). The corresponding vQTL lead variants were rs28633797, an intronic variant of *RHCE*, and rs71186552, an intronic variant of *PACSIN2*. The African, East Asian, and South Asian ancestry-specific analyses were not well-powered likely due to the limited sample sizes (**Supplementary Table S1**). Only one QTL in *GC* was identified in the African and South Asian ancestry populations, respectively, while no vQTLs were identified in these non-European ancestry populations (**Supplementary Figures S2-S4**). Q-Q plots suggested that these analyses were well-controlled for potential confounding factors (**Supplementary Figure S5**).

**Figure 1.**
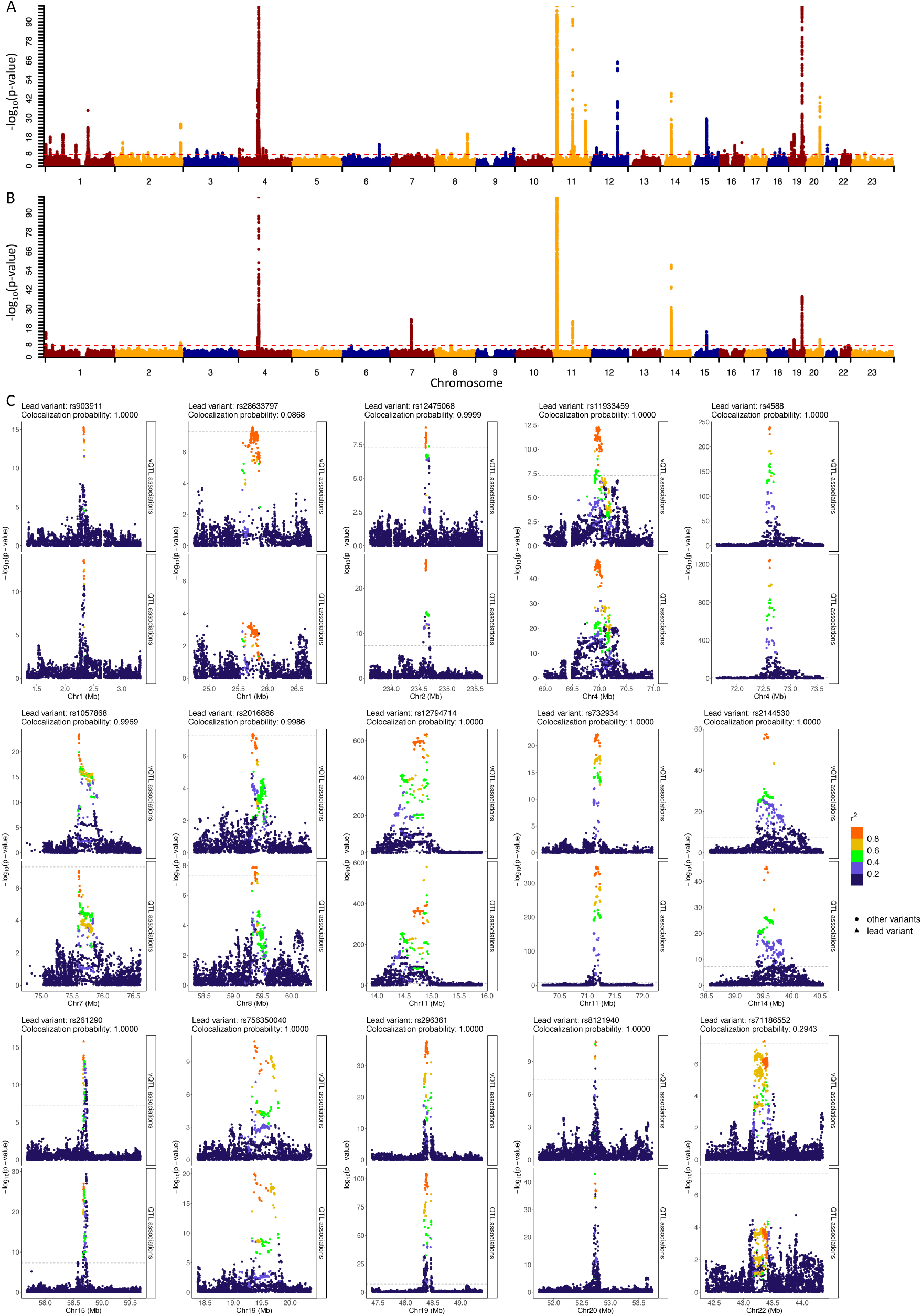
Identification of variance quantitative trait loci (vQTLs) for serum 25-hydroxy vitamin D (25OHD) levels. Genetic associations with (A) trait mean and (B) trait variance are illustrated in Manhattan plots. P-values <1.0×10^-100^ are set to 1.0×10^-100^ for visualization purposes. (C) Locus-zoom plots comparing the genetic associations with the trait mean (QTL associations) and the trait variance (vQTL associations). The vQTL lead variants and colocalization probabilities are indicated. Genetic variants located within a ±500kb window centered around each vQTL lead variant are plotted with their significance and colored by the magnitude of correlation (linkage disequilibrium, LD r^2^) with the corresponding lead variant. Red dashed lines indicate the genome-wide significance threshold (p-value <5.0×10^-8^).

The European ancestry-specific vQTLs withstood multiple sensitivity analyses. For each vQTL lead variant, as illustrated using conditional quantile regression, the trend of varying genetic effects was not sensitive to inverse rank normal transformation of serum 25OHD concentrations (**Supplementary Figures S6-S7** and **Supplementary Table S3**). All 19 vQTL lead variants had a p-value <1.0×10^-5^ with alternative methods for homogeneity of variance test, including 17 demonstrating genome-wide significance (p-value <5.0×10^-8^) using the mean-based Levene’s test, 14 using the median-based Levene’s test, and 16 using the Fligner-Killeen test (**Supplementary Table S4**). Furthermore, after including 12,119 additional European ancestry individuals who self-reported being on vitamin D supplementation, no additional vQTLs were identified (**Supplementary Figure S8**). Meanwhile, 17 of the 19 originally identified lead variants demonstrated genome-wide significance, while the remaining two lead variants were suggestively significant (rs2016886, p-value = 1.2×10^-7^; rs1792581, p-value = 7.5×10^-8^; **Supplementary Table S5**).

### Gene-environment interaction effects identified in variance quantitative trait loci

For each of the 19 vQTL lead variants, we tested for interaction effects with each of the 18 candidate environmental factors, encompassing the season of measurement, two non-modifiable biological factors (age and sex), two adiposity measures (BMI and WHR), nine lifestyle factors (smoking status, alcohol intake, time spent outdoors, and the first six principal components of food intake variables; **Supplementary Figure S9A**), as well as four kidney or liver function biomarkers (eGFR, ALT, AST, and GGT).

As a result, we identified 32 significant variant-environmental factor interaction effects with an FDR <0.05 accounting for a total of 342 tests, including 18 interaction effects having a Bonferroni-corrected p-value <0.05 (**Figure 2A** and **Supplementary Table S6**). These 32 interaction effects involved 10 of the 19 vQTL lead variants. With inverse rank normal transformed serum 25OHD concentrations as the outcome, most of these interaction effects remained directionally consistent and nominally significant, except that the interaction between rs732934 (eQTL and sQTL of *DHCR7*) and season of measurement was no longer significant (p-value = 0.46; **Supplementary Table S6**).

**Figure 2.**
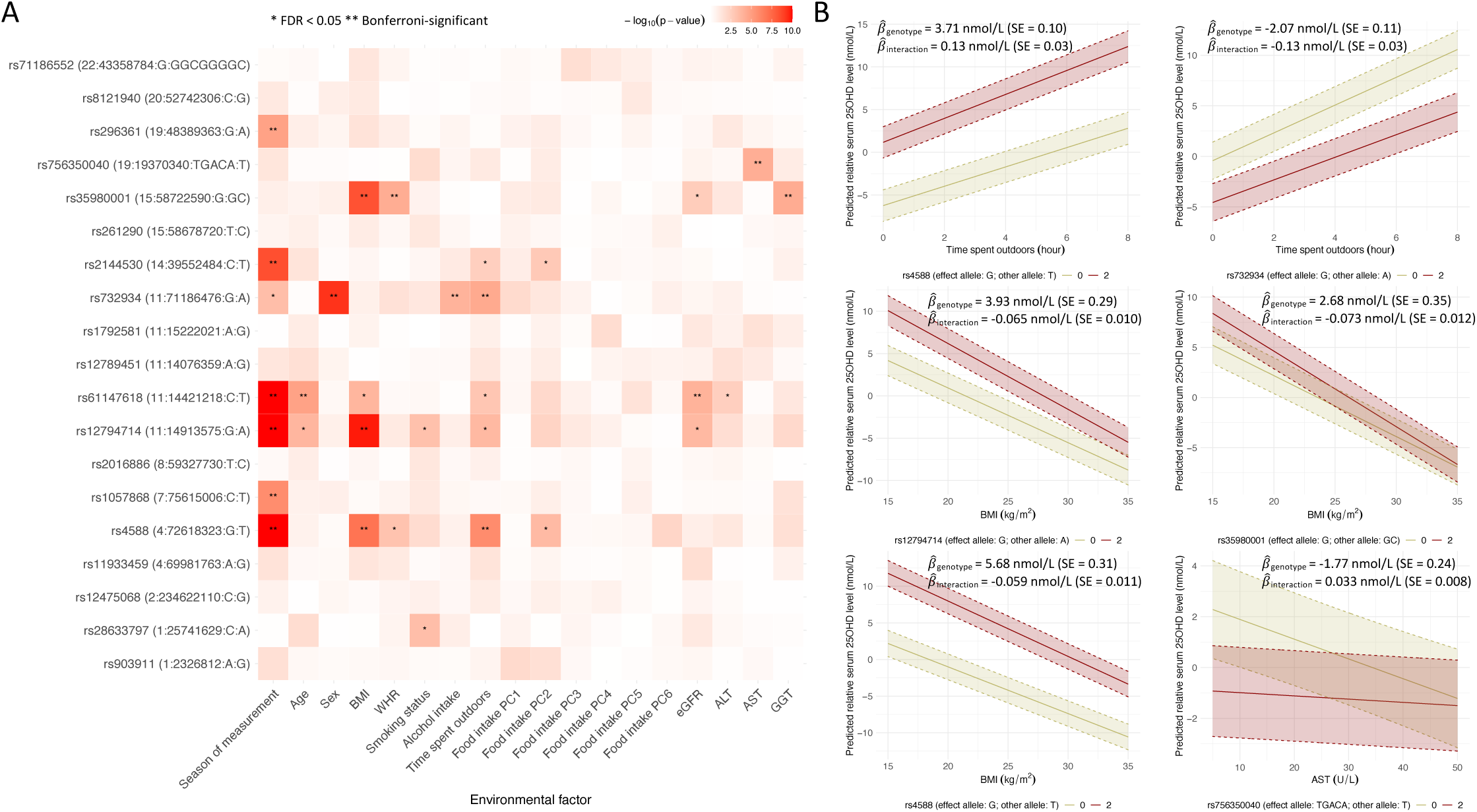
Identification of gene-environment interaction effects affecting vitamin D status. (A) Illustration of pairwise interaction effects between each vQTL lead variant and each of the 18 candidate environmental factors based on single-interaction models. P-values <1.0×10^-10^ are set to 1.0×10^-10^ for visualization purposes. Results based on multiple-interactions models are available in Supplementary Figure S8 and Supplementary Table S7. (B) Illustration of selected gene-environment interaction effects. Predicted relative serum 25-hydroxy vitamin D (25OHD) levels (solid lines) and 95% prediction intervals (dashed lines) are plotted as a function of the genotype of the genetic variant and the environmental factor that demonstrated interaction effects. Baseline 25OHD levels were obtained with two hours of time spent outdoors, a BMI of 25 kg/m^2^, and a serum AST level of 20 U/L, respectively. The estimated main effect of the genotype 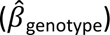 and the interaction effect 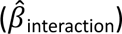 are indicated for selected vQTL lead variants. The interaction effects corresponding to a 1-hour increase in time spent outdoors, a 1 kg/m^2^ increase in BMI, or a 1 U/L increase in serum AST levels.

As expected, season of measurement exhibited interaction effects with seven vQTL lead variants (**Figure 2A**). Five of these vQTL lead variants also showed significant interaction effects with time spent outdoors (**Figure 2A**). Genetic effects of these five vQTL lead variants were stronger with increased time spent outdoors (**Figure 2B** and **Supplementary Table S6**). For instance, the effect allele of rs4588 (missense variant of *GC*) had an estimated main effect of 3.71 nmol/L (SE = 0.10 nmol/L) on serum 25OHD concentrations with an estimated interaction effect of 0.13 nmol/L (SE = 0.03 nmol/L) per 1-hour increase in time spent outdoors, while the effect allele of rs732934 (eQTL and sQTL of *DHCR7*) had an estimated main effect of -2.07 nmol/L (SE = 0.11 nmol/L) with an estimated interaction effect of -0.13 nmol/L (SE = 0.03 nmol/L; **Figure 2B** and **Supplementary Table S6**). These variant-time spent outdoors interaction effects were consistent in multiple-interactions models that included variant-season of measurement interaction effects (**Supplementary Figure S10** and **Supplementary Table S7**). In contrast, two variant-age interaction effects were attenuated in multiple-interactions models (**Supplementary Figure S10** and **Supplementary Table S7**).

Multiple interaction effects were detected between vQTL lead variants and adiposity measures and lifestyle factors, where the strongest interaction effects involved BMI (**Figure 2A**). Genetic effects of four vQTL lead variants, rs12794714 (synonymous variant and eQTL of *CYP2R1*), rs35980001 (intronic variant of *LIPC* and *ALDH1A2*), rs4588 (missense variant of *GC*), and rs61147618 (intergenic variant within 500kb of *CYP2R1*), weakened with increased BMI (**Figure 2B** and **Supplementary Table S6**). These interaction effects were consistent in multiple-interactions models (**Supplementary Figure S10** and **Supplementary Table S7**). Two of these four vQTL lead variants also showed interaction effects with WHR (**Figure 2A**). However, the variant-WHR interaction effects were attenuated in the respective multiple-interactions models (**Supplementary Figure S10** and **Supplementary Table S7**). Furthermore, smoking status demonstrated consistent interaction effects with rs12794714 (synonymous variant and eQTL of *CYP2R1*) and rs28633797 (intronic variant of *RHCE*), while the rs732934 (eQTL and sQTL of *DHCR7*)-alcohol intake interaction effect was attenuated in the multiple-interactions model (**Supplementary Figure S10** and **Supplementary Table S7**).

Of the first six principal components of food intake, the second principal component was consistently correlated with potential dietary sources of vitamin D (**Supplementary Figure S9B**).

This principal component demonstrated interaction effects with rs4588 (missense variant of *GC*) and rs2144530 (intronic variant of *SEC23A*; **Figure 2A**), which were consistent in multiple-interactions models (**Supplementary Figure S10** and **Supplementary Table S7**).

Among the investigated biomarkers, serum AST concentrations demonstrated a significant interaction effect with rs756350040, an intronic variant of *HAPLN4* (**Figure 2A** and **B** and **Supplementary Table S6**). At other vQTLs, variant-biomarker interaction effects involving eGFR, ALT, or GGT were attenuated in multiple-interactions models (**Supplementary Figure S10** and **Supplementary Table S7**).

### Sex-differentiated genetic effects underlying *DHCR7*

In the European ancestry population, males on average had higher concentrations of serum 25OHD compared to females (mean = 49.6 nmol/L, SD = 21.0 nmol/L in males vs. mean = 49.3 nmol/L, SD = 20.8 nmol/L in females; t-test p-value = 2.4×10^-7^). Variant-sex interaction effect was detected only at rs732934 (**Figure 2A**), which was consistent in the multiple-interactions model (**Supplementary Figure S10** and **Supplementary Table S7**). The effect allele of rs732934 had an estimated effect of -2.04 nmol/L (SE = 0.08 nmol/L; p-value = 1.0×10^-128^) on serum 25OHD concentrations in females and -2.79 nmol/L (SE = 0.09 nmol/L; p-value = 1.7×10^-232^) in males (**Figure 3A**). This variant is a known eQTL and sQTL variant for *DHCR7* in both sun-exposed lower leg skin and not-sun-exposed suprapubic skin tissues in GTEx (**Supplementary Table S8**). Notably, the 25OHD-decreasing allele of rs732934 was associated with increased overall DHCR7 transcript abundance as well as an increased excision rate of an intron (chromosome 11:71444119-71444855, genome assembly GRCh38.p14; **Supplementary Table S8**). The eQTL effect appeared to be stronger in males (a 0.18 SD increase in normalized expression levels; SE = 0.05; p-value = 3.8×10^-4^) than in females (a 0.12 SD increase in normalized expression levels; SE = 0.05; p-value = 2.6×10^-2^) in sun-exposed lower leg skin tissues, although this difference was not significant (**Supplementary Table S8**).

**Figure 3.**
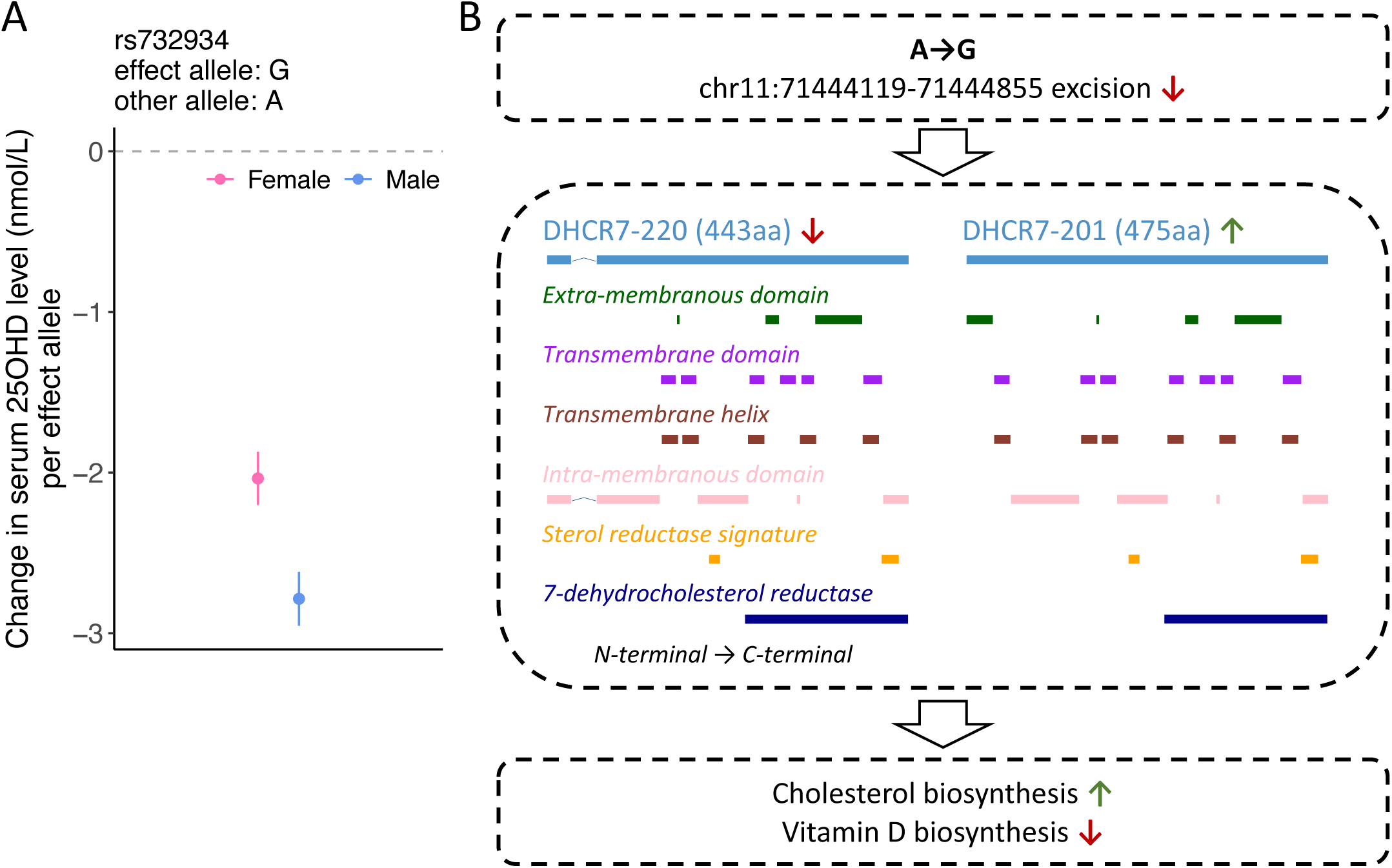
Genetic effects mapping to DHCR7. (A) Illustration of the sex-differentiated genetic effects of rs732934 on serum 25-hydroxy vitamin D (25OHD) levels. Error bars indicate 95% confidence intervals. (B) Protein isoforms resulting from alternative splicing associated with rs732934. The 25OHD-decreasing allele has been associated with decreased proportion of DHCR7-220 and increased proportion of DHCR7-201 in skin tissues. DHCR7-201 contains an additional transmembrane domain and an additional extra-membranous domain at the N-terminal compared to DHCR7-220. Amino acid sequences and annotations of structural and functional domains are available in Supplementary Table S9.

Interestingly, alternative splicing of *DHCR7* leads to two protein isoforms: DHCR7-201 (Ensembl transcript ID: ENST00000355527.8) and DHCR7-220 (Ensembl transcript ID: ENST00000690257.1). The 25OHD-decreasing allele of rs732934 decreases the proportion of DHCR7-220 while increasing the proportion of DHCR7-201, which has a cytosine-to-tryptophan mutation (C33W) as well as 32 additional amino acids compared to the former (**Supplementary Table S9**). DHCR7-201 is predicted to have an additional transmembrane domain and extra-membranous domain at the N-terminal compared to the former, although both isoforms share the same functional domain of 7-dehydrocholesterol reductase (**Figure 3B** and **Supplementary Table S9**). Importantly, overall expression levels of *DHCR7* were higher in males than in females in skin (LFSR^35^ = 1.9×10^-6^ in sun-exposed lower leg skin and 5.2×10^-4^ in not-sun-exposed suprapubic skin), as well as in several other tissues (**Supplementary Figure S11**).

## Discussion

Vitamin D deficiency is a global health challenge that impacts various populations. In this study, we leveraged the UK Biobank to characterize gene-environment interaction effects on vitamin D status through the identification of vQTLs. In addition to season of measurement, we found that several modifiable risk factors, such as time spent outdoors and BMI, demonstrated interaction effects with vQTL lead variants on serum 25OHD concentrations. Furthermore, transcript- and protein-level evidence suggests that the identified sex-differentiated genetic effects might act through sex-differential expression of DHCR7 isoforms in skin tissues.

vQTLs identified in this study are consistent with the literature. Specifically, all vQTLs identified in a previous study using the median-based Levene’s test in the UK Biobank European ancestry population were confirmed in this study^7^. Using QUAIL, we detected three additional vQTLs (lead variants: rs12475068, rs28633797, and rs71186552). rs12475068 is located in the *UGT1A* locus and has functional impacts on several genes that encode various UGT1A enzymes (**Supplementary Table S2**). This variant is close to a microsatellite variant that may have causal effects on Gilbert’s syndrome (OMIM 143500)^38^. These enzymes catalyze the glucuronidation of vitamin D metabolites to produce more water-soluble glucuronides, likely contributing to the maintenance of vitamin D homeostasis^39^. This variant also had a strong mean effect on serum 25OHD concentrations. In contrast, rs28633797 and rs71186552 only demonstrated variance effects. Interestingly, these two variants have been associated with the amount and morphology of platelets (rs28633797 and rs71186552) and erythrocytes (rs28633797; **Supplementary Table S2**). These associations may reflect the anti-inflammatory and immunomodulatory effects of vitamin D^2^, as well as its involvement in hematopoiesis^40^. However, since both variants demonstrated eQTL or sQTL effects on multiple neighboring genes, further investigations are needed to pinpoint the target genes.

Our gene-environment interaction analyses identified interaction effects involving several modifiable risk factors for vitamin D deficiency while confirming known gene-season of measurement interaction effects. Despite correlation between the candidate environmental factors, interaction effects involving either time spent outdoors or BMI at specific vQTLs were not attenuated in multiple-interactions models. As expected, as a direct indicator of sunlight exposure, time spent outdoors demonstrated interaction effects with most of the vQTL lead variants that also interacted with season of measurement. At these vQTL lead variants, increased time spent outdoors amplified the effects of 25OHD-increasing alleles, reinforcing the critical role of sunlight exposure in vitamin D biosynthesis.

Among environmental factors unrelated to sunlight exposure, BMI demonstrated the strongest gene-environment interaction effects, with increased BMI mitigating the genetic effects. Interestingly, if having 25OHD-increasing alleles can be analogized to lifetime exposure to low-dose vitamin D supplementation, these results align with a recent randomized clinical trial showing that the effect of vitamin D supplementation on serum 25OHD is weaker amongst individuals with a higher BMI^41^. This complies with the hypothesis that, as a fat-soluble vitamin, the amount of vitamin D in its circulating form can be reduced with increased deposition in body fat compartments^42,43^. In contrast, the vQTL variant-WHR interaction effects were attenuated when adjusting for BMI, suggesting that overall body fat may play a more important role than fat distribution in regulating vitamin D status.

Although essential reactions for producing biologically active vitamin D occur in the liver and kidneys, most interaction effects involving liver or kidney function biomarkers were attenuated in the multiple-interactions models, except for AST. Nonetheless, since elevated serum AST concentrations are also associated with tissue damage in other organs^44^, this may not be specifically interpreted as a gene-liver function interaction effect. It should be noted that the majority of the study population had these biomarkers within normal ranges^45^. Therefore, it remains possible that potential interactions affect individuals with impaired kidney or liver function. Along with potential interaction effects involving smoking status and dietary intake of vitamin D (the second principal component of food intake) that were not Bonferroni-significant, these findings require further investigation in future studies.

Gene-sex interaction effects were identified at rs732934, which influences the expression of *DHCR7*. Gene expression profiles in GTEx indicate that the sex-differentiated genetic effects may be mediated by sex-differential gene expression. In skin tissues, DHCR7 catalyzes the final step of the Kandutsch-Russell pathway^46^ and is a switch between cholesterol biosynthesis and vitamin D biosynthesis^47^, since both reactions utilize the same substrate, 7-dehydrocholesterol. Alternative splicing associated with the 25OHD-decreasing allele of rs732934 can lead to increased expression of DHCR7-201. Despite the preserved functional domain, the structural difference between DHCR7-201 and DHCR7-220 at the N-terminal might alter the stability of the enzyme when localizing to the endoplasmic reticulum membrane and nuclear outer membrane^48^. These hypotheses warrant further functional characterization of DHCR7 isoforms and may potentially identify new molecular targets for managing vitamin D deficiency.

This study has important limitations. First, all vQTLs were identified in the European ancestry population. It is unclear whether the European ancestry-specific variance effects and gene-environment interaction effects also influence vitamin D status in populations of non-European ancestries due to their limited sample sizes in the UK Biobank. Second, our results have not been validated in other cohort studies. The UK Biobank participants were middle-aged or older individuals living in the United Kingdom at the time of recruitment, and were, on average, healthier and less socioeconomically deprived than the non-participants^45^. The identified gene-environment interaction effects may not be generalizable to populations with different characteristics. Third, no significant interaction effects were detected at nine of the 19 vQTL lead variants. This may suggest the existence of unmodeled gene-environment interaction effects, epistatic effects, or other complex regulatory mechanisms. However, replicating known and identifying additional interaction effects require sufficient statistical power, as the magnitudes of the interaction effects are often smaller than those of the main effects. Additional investigations, possibly in a longitudinal setting, are strongly warranted in diverse populations with increased sample sizes. Furthermore, gene-environment interaction analyses can be biased by unmeasured confounders or reverse causation. While previous Mendelian randomization studies indicated that lower serum 25OHD concentrations are more likely to be the consequence, rather than the cause, of various risk factors such as increased BMI^49^, causal interpretations will only be justified with rigorous functional follow-up studies and triangulation of multiple lines of evidence. Last, although our study did not identify additional vQTLs after including individuals who self-reported being on vitamin D supplementation, precise measurement of vitamin D supplementation and other medications may better characterize gene-supplementation interaction effects, which could have important clinical and public health implications.

In summary, this work has expanded known gene-environment interactions that affect vitamin D status. These findings may help better understand the etiology of vitamin D deficiency, refine public health guidelines on healthy lifestyle, and possibly tailor interventions to prevent vitamin D deficiency-related health issues for at-risk individuals.

## Acknowledgements

This research has been conducted using the UK Biobank Resource under Application Number 64875. UK Biobank has approval from the North West Multi-centre Research Ethics Committee (MREC) as a Research Tissue Bank (RTB) approval. This approval means that researchers do not require separate ethical clearance and can operate under the RTB approval (https://www.ukbiobank.ac.uk/learn-more-about-uk-biobank/about-us/ethics).

## Author Contributions

T.L.: conceptualization, investigation, formal analysis, visualization, and writing original draft; T.L., L.S., A.D.P.: resources; T.L., W.Z., L.S., A.D.P.: methodology; all authors contributed to interpretation of results, reviewed and edited the manuscript critically, and approved the final manuscript.

## Data Availability

Full summary statistics for both vQTL and QTL analyses are available at https://figshare.com/s/a6c1d92d9cc765e0d58b.

**Supplementary Tables are available at https://figshare.com/s/82f05d4c830dc4af50ac**

