## Supplementary material for "Improved characterization of gene-environment interactions for vitamin D through variance quantitative trait loci": Supp Figures

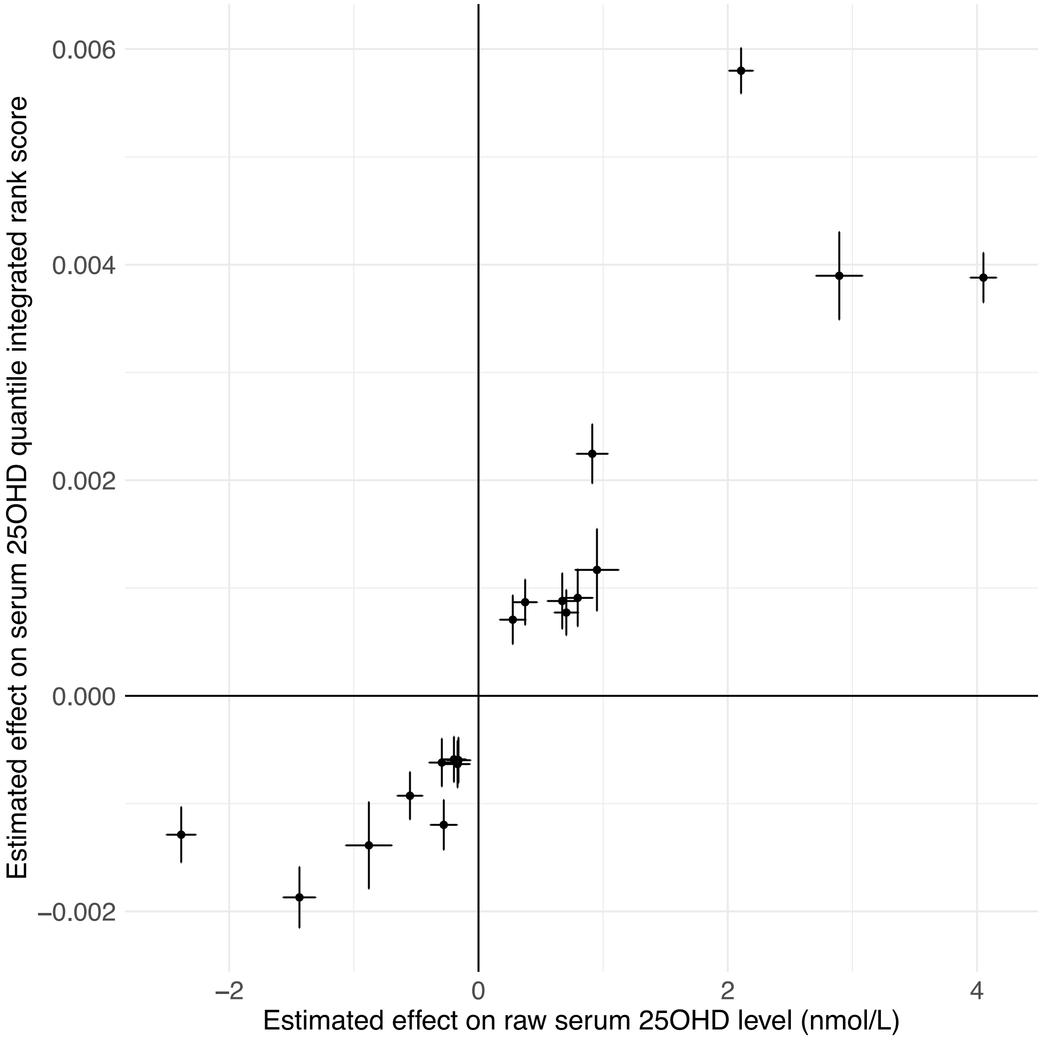


**Supplementary Figure S1.** Comparison of mean effects (QTL) and variance effects (vQTL) on serum 25-hydroxy vitamin D (25OHD) levels in the European ancestry population. Each dot represents one vQTL lead variant. Error bars indicate 95% confidence intervals. Spearman correlation = 0.96 (p-value = $7.5 \times 10^{-6}$).

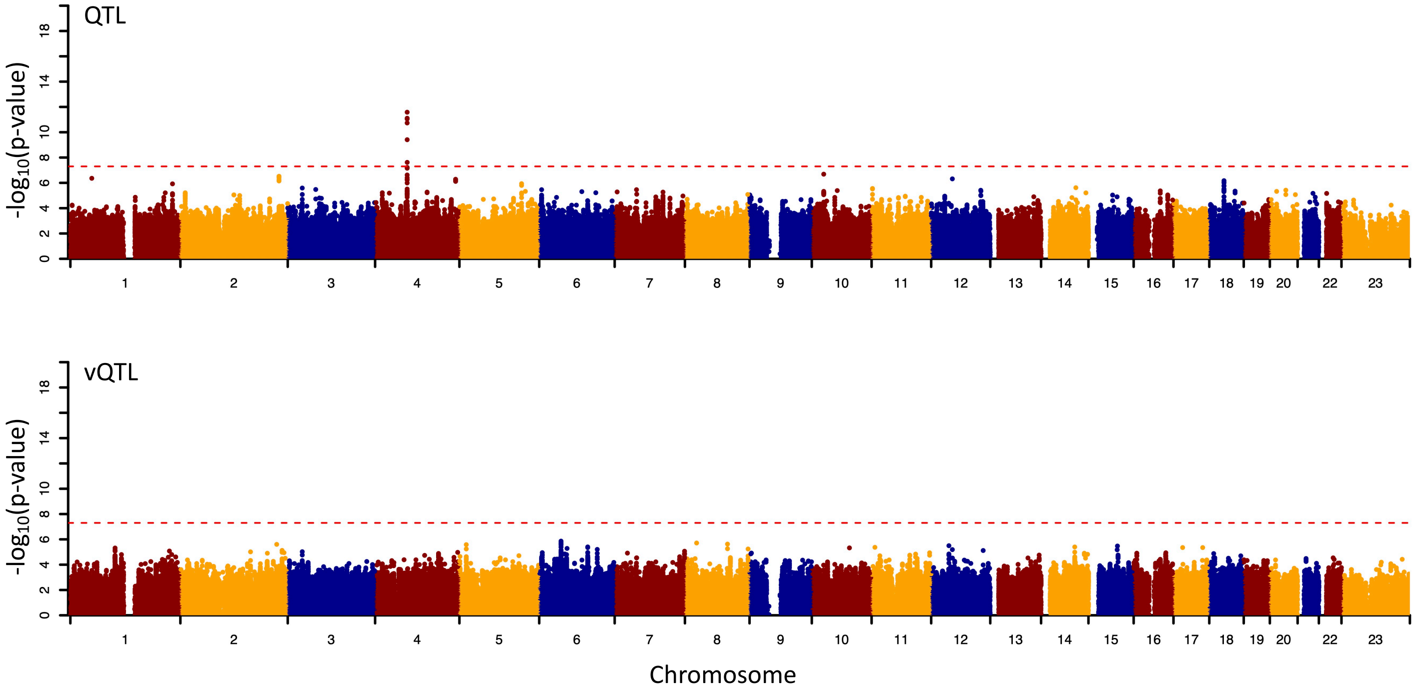


**Supplementary Figure S2.** Manhattan plots showing genetic associations with trait mean (QTL) and trait variance (vQTL) of serum 25-hydroxy vitamin D (25OHD) levels in the African ancestry population. Red dashed lines indicate the genome-wide significance threshold (p-value $<5.0\text{x}10^{-8}$).

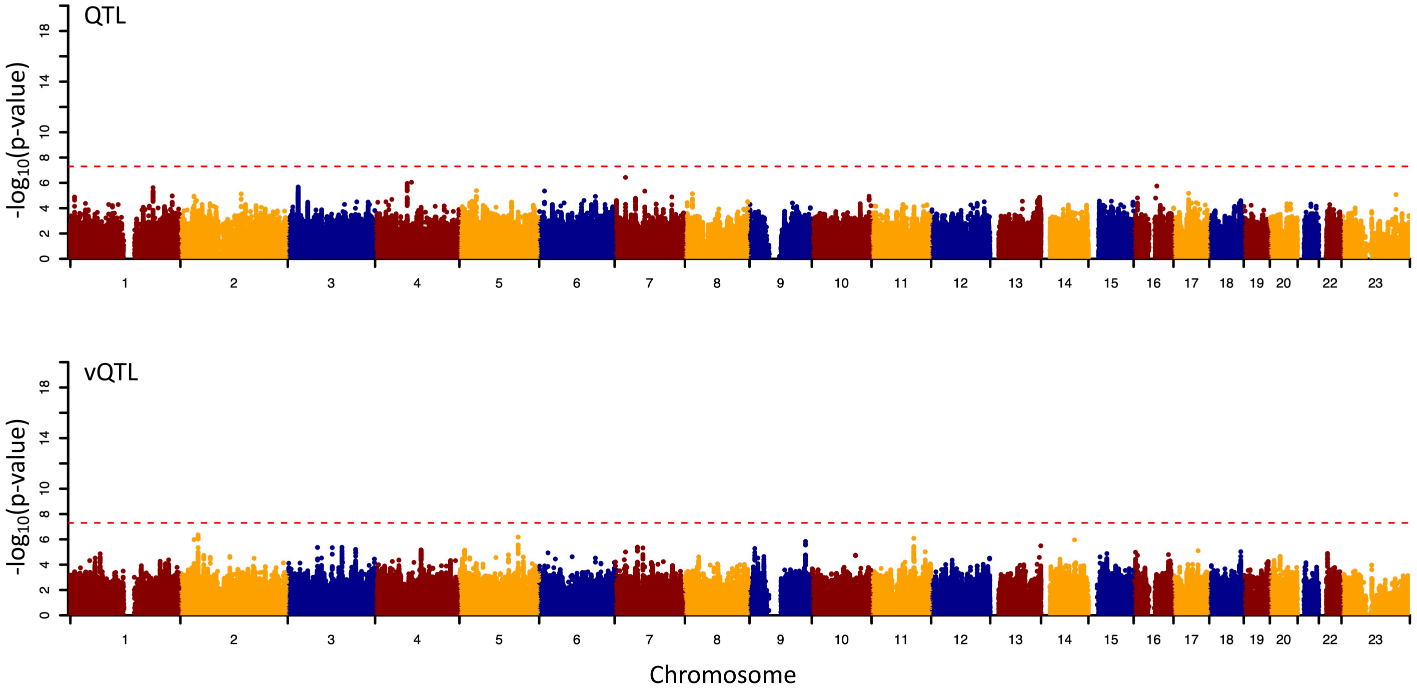


**Supplementary Figure S3.** Manhattan plots showing genetic associations with trait mean (QTL) and trait variance (vQTL) of serum 25-hydroxy vitamin D (25OHD) levels in the East Asian ancestry population. Red dashed lines indicate the genome-wide significance threshold (p-value $<5.0\text{x}10^{-8}$).

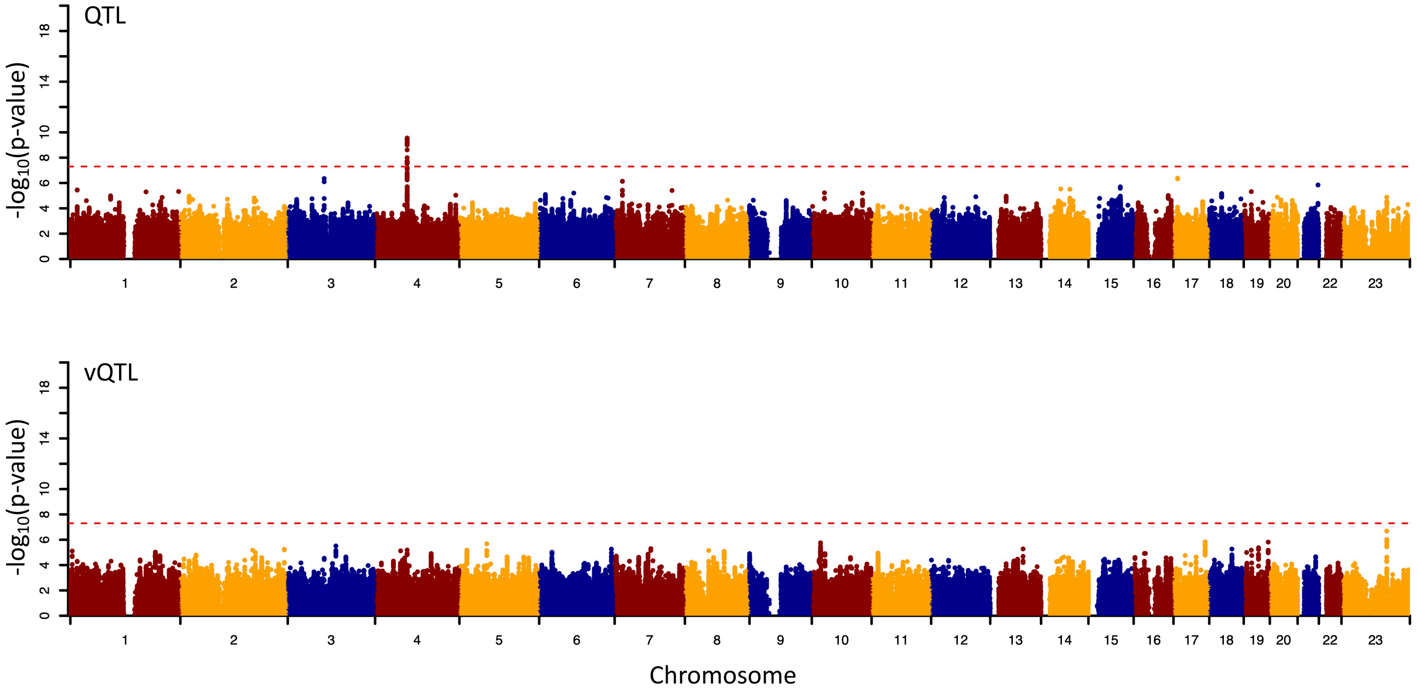


**Supplementary Figure S4.** Manhattan plots showing genetic associations with trait mean (QTL) and trait variance (vQTL) of serum 25-hydroxy vitamin D (25OHD) levels in the South Asian ancestry population. Red dashed lines indicate the genome-wide significance threshold (p-value $<5.0x10^{-8}$).

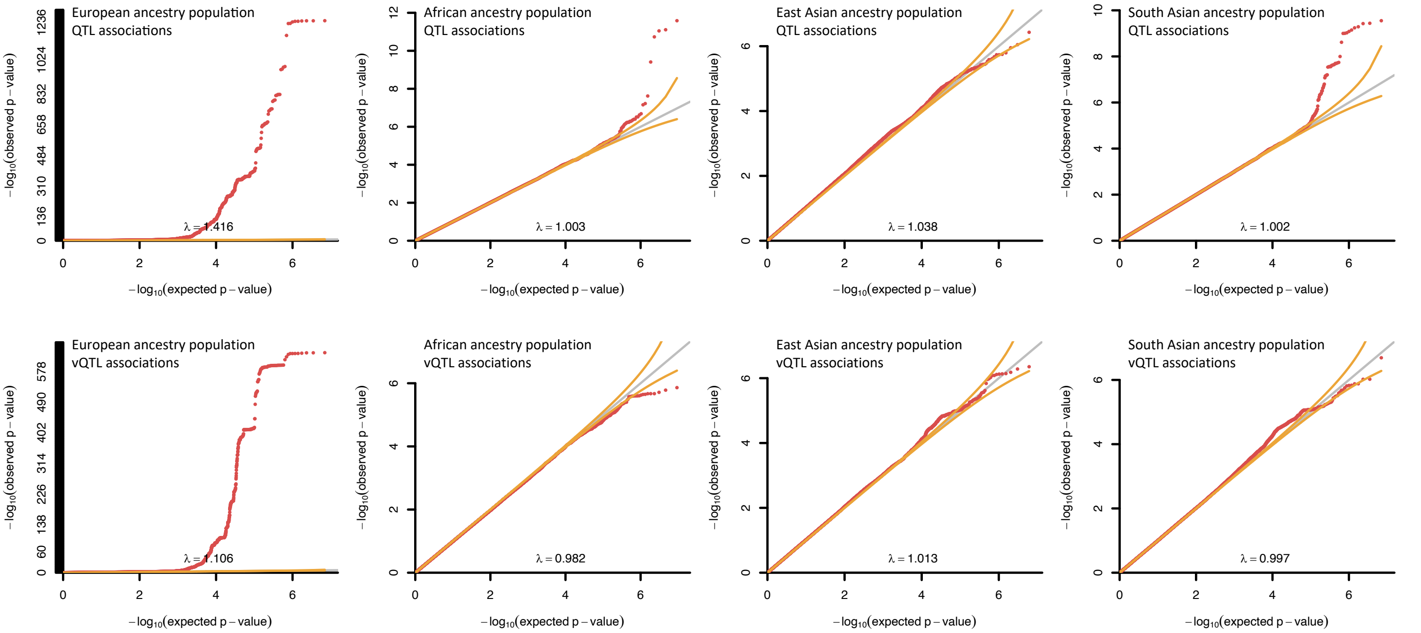


**Supplementary Figure S5.** Quantile-quantile plots illustrating the distributions of p-values in QTL and vQTL analyses. The genomic inflation factors ($\lambda$) are indicated. Red dots indicate p-values of the genetic variants across the genome. Grey lines have a slope of 1 with orange lines indicating the 95% confidence intervals.

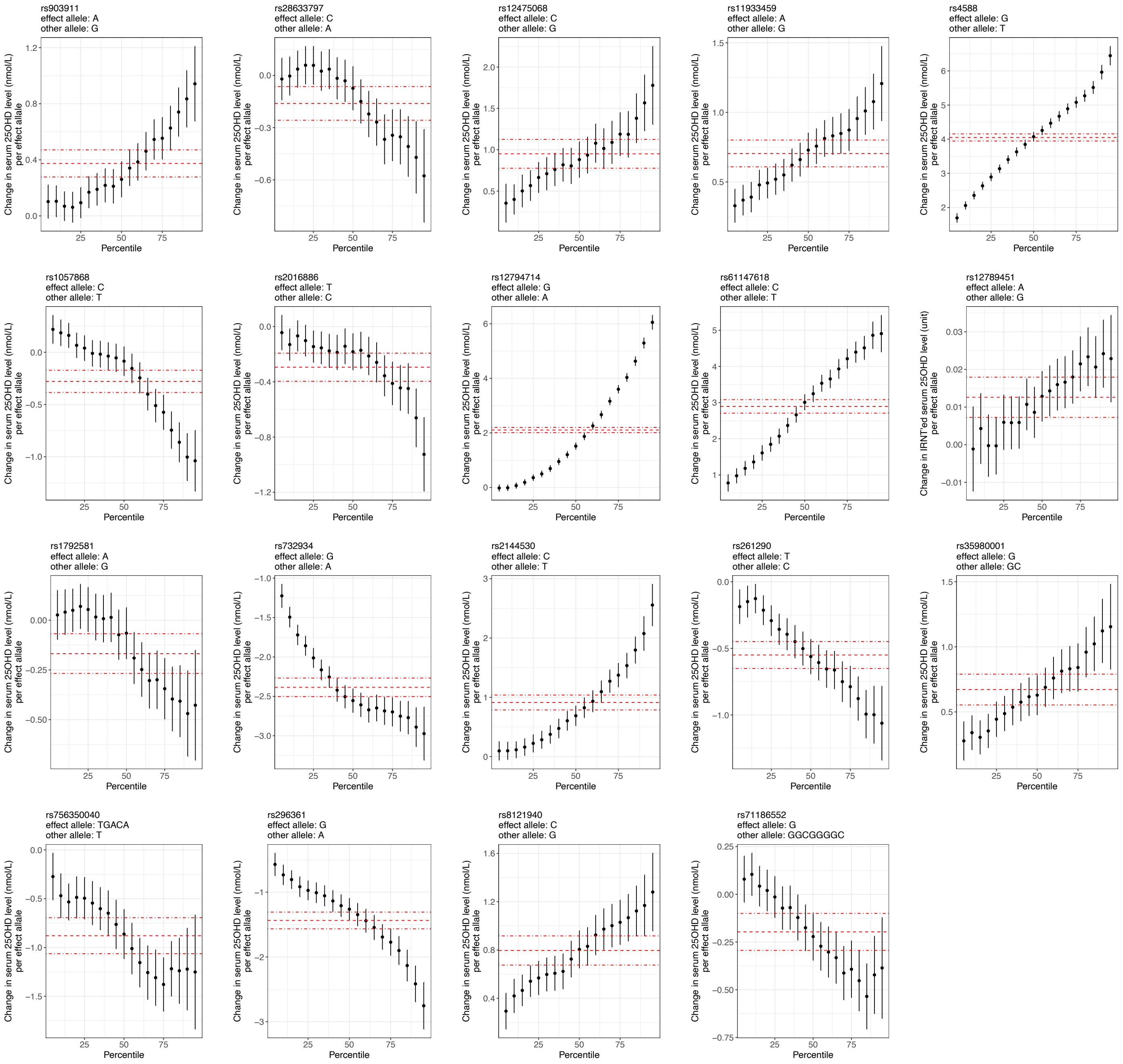


**Supplementary Figure S6.** Estimated genetic effects of vQTL lead variants on raw serum 25-hydroxy vitamin D (25OHD) levels using conditional quantile regression. Dots represent conditional quantile regression estimates obtained at a series of quantiles: 0.05, 0.10, 0.15, 0.20, 0.25, 0.30, 0.35, 0.40, 0.45, 0.50, 0.55, 0.60, 0.65, 0.70, 0.75, 0.80, 0.85, 0.90, and 0.95. Error bars represent 95% confidence intervals. Red dashed lines indicate the mean effect estimates in GWAS with dash-dotted lines indicating the 95% confidence intervals.

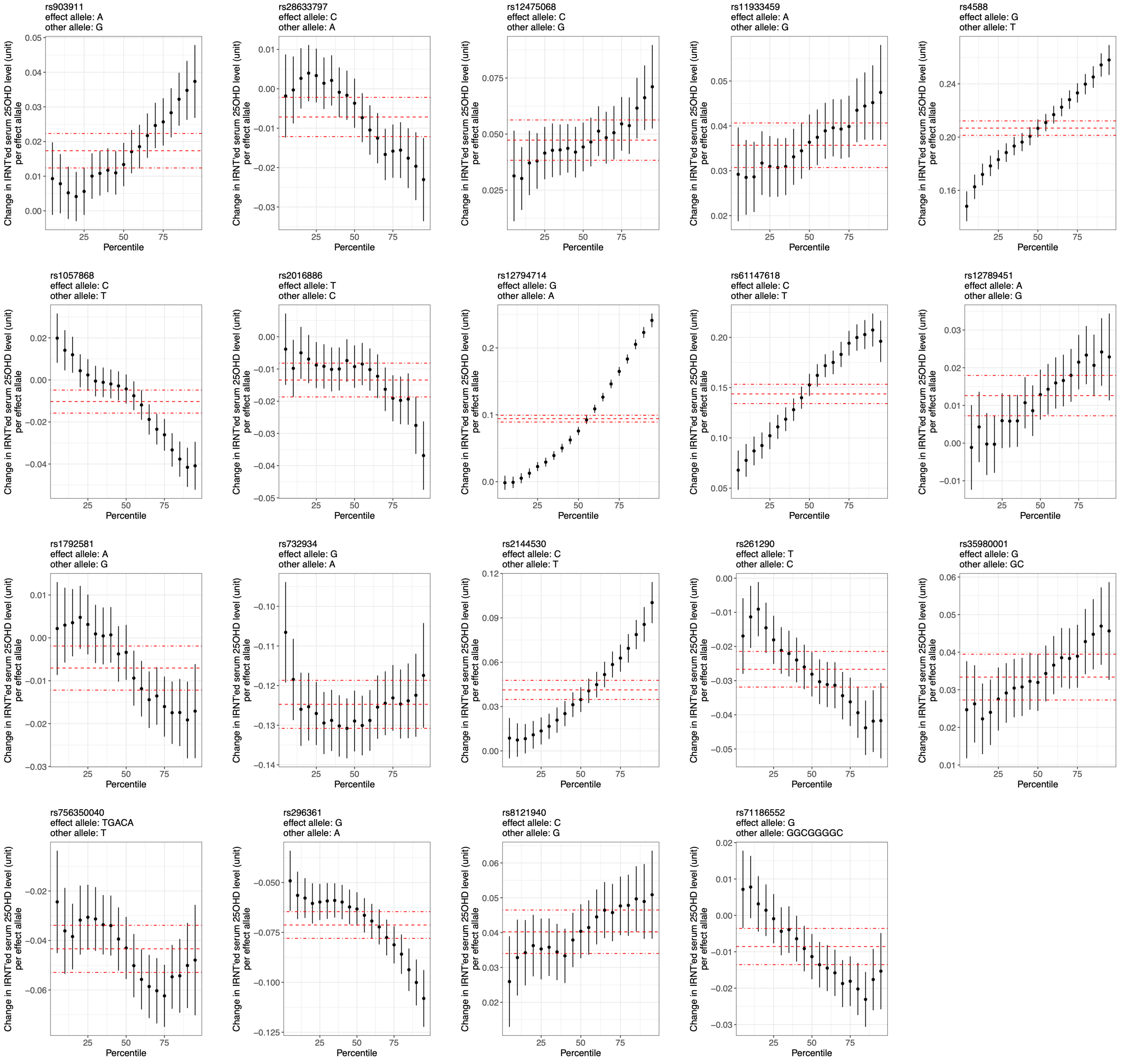


**Supplementary Figure S7.** Estimated genetic effects of vQTL lead variants on inverse normal transformed (IRNT'ed) serum 25-hydroxy vitamin D (25OHD) levels using conditional quantile regression. Dots represent conditional quantile regression estimates obtained at a series of quantiles: 0.05, 0.10, 0.15, 0.20, 0.25, 0.30, 0.35, 0.40, 0.45, 0.50, 0.55, 0.60, 0.65, 0.70, 0.75, 0.80, 0.85, 0.90, and 0.95. Error bars represent 95% confidence intervals. Red dashed lines indicate the mean effect estimates in GWAS with dash-dotted lines indicating the 95% confidence intervals.

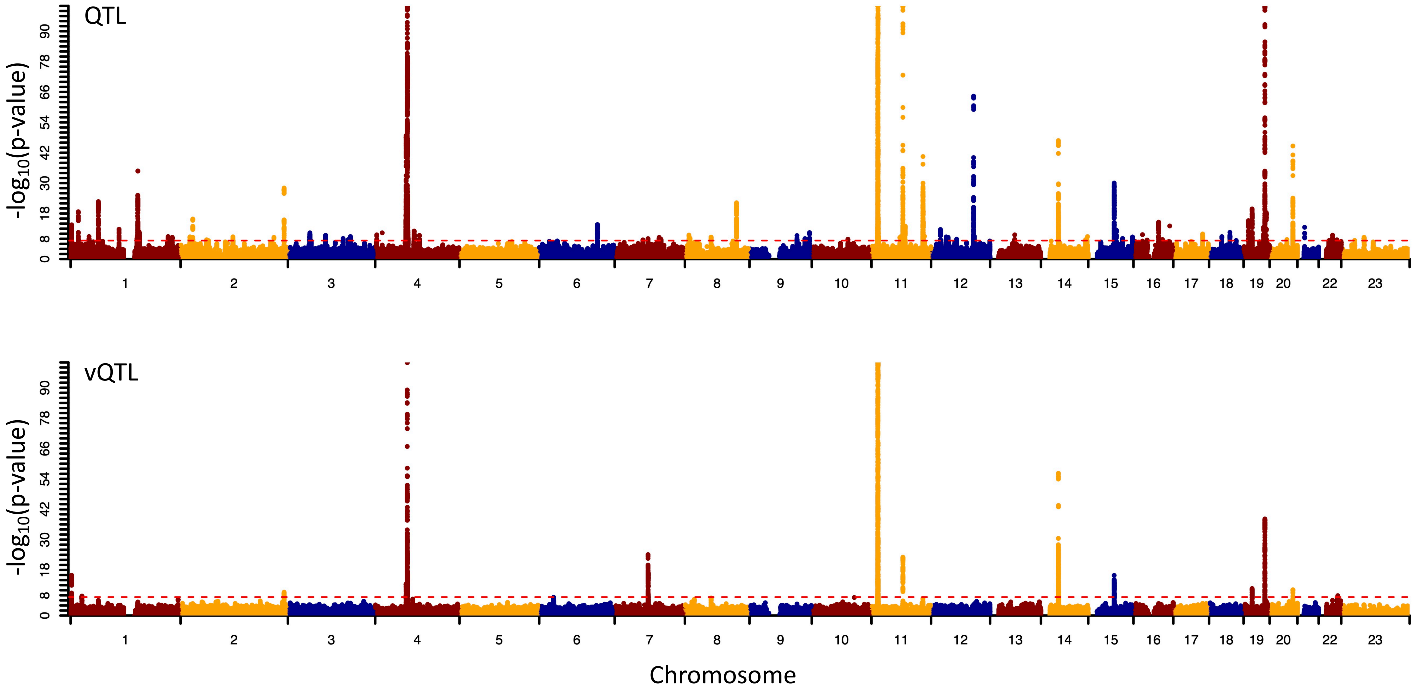


**Supplementary Figure S8.** Manhattan plots showing genetic associations with trait mean (QTL) and trait variance (vQTL) of serum 25-hydroxy vitamin D (25OHD) levels in the European ancestry population including 12,119 individuals who self-reported being on vitamin D supplementation. Red dashed lines indicate the genome-wide significance threshold (p-value $<5.0\times10^{-8}$).

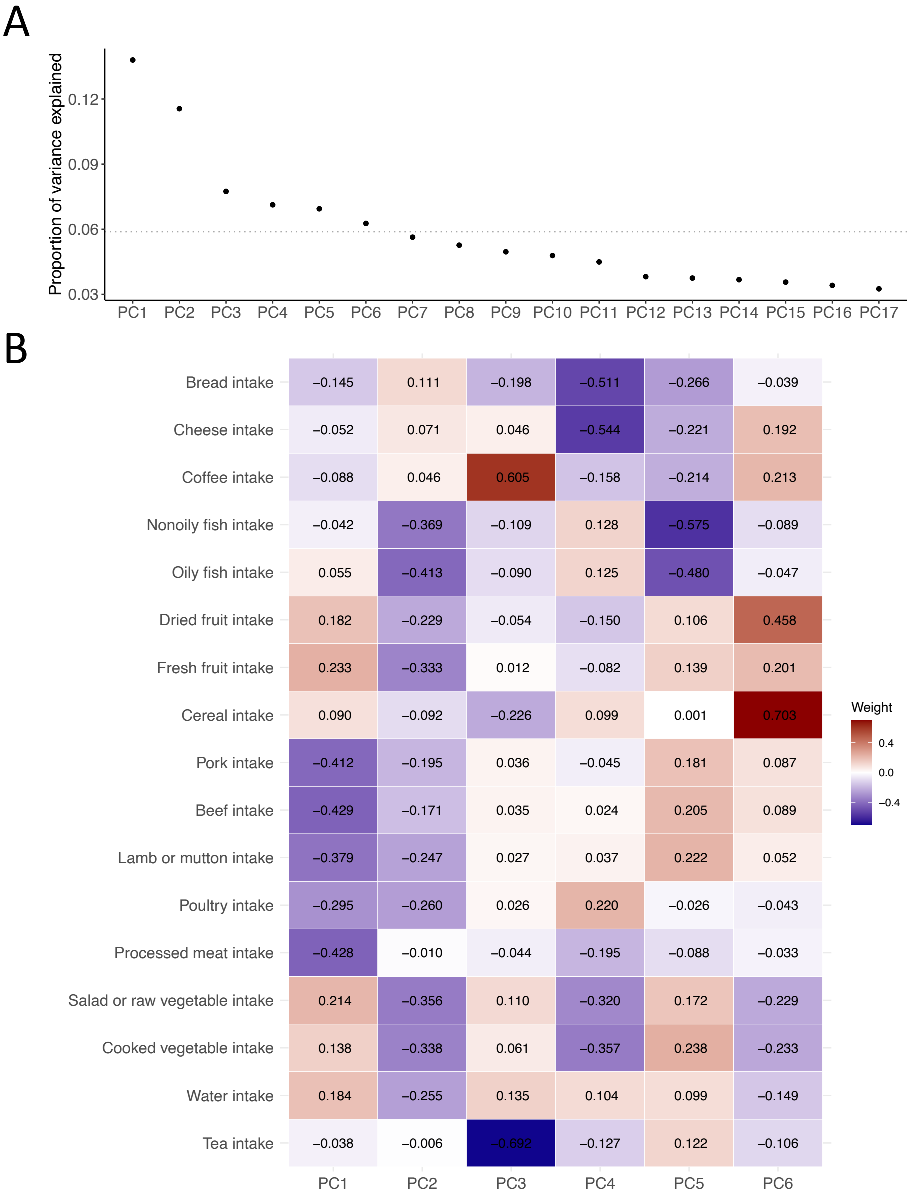


**Supplementary Figure S9.** Principal component analysis of food intake variables. (A) Proportion of variance explained by each of the 17 principal components. The first six principal components captured more than 1/17 of the total variance (indicated by the dotted line) and were included in gene-environment interaction analyses. (B) Weight of each food intake variable in each of the first six principal components. The second principal component was negatively associated with potential dietary sources of vitamin D, such as oily fish and animal-source foods which may include liver. Food intake variables are ordered based on hierarchical clustering.

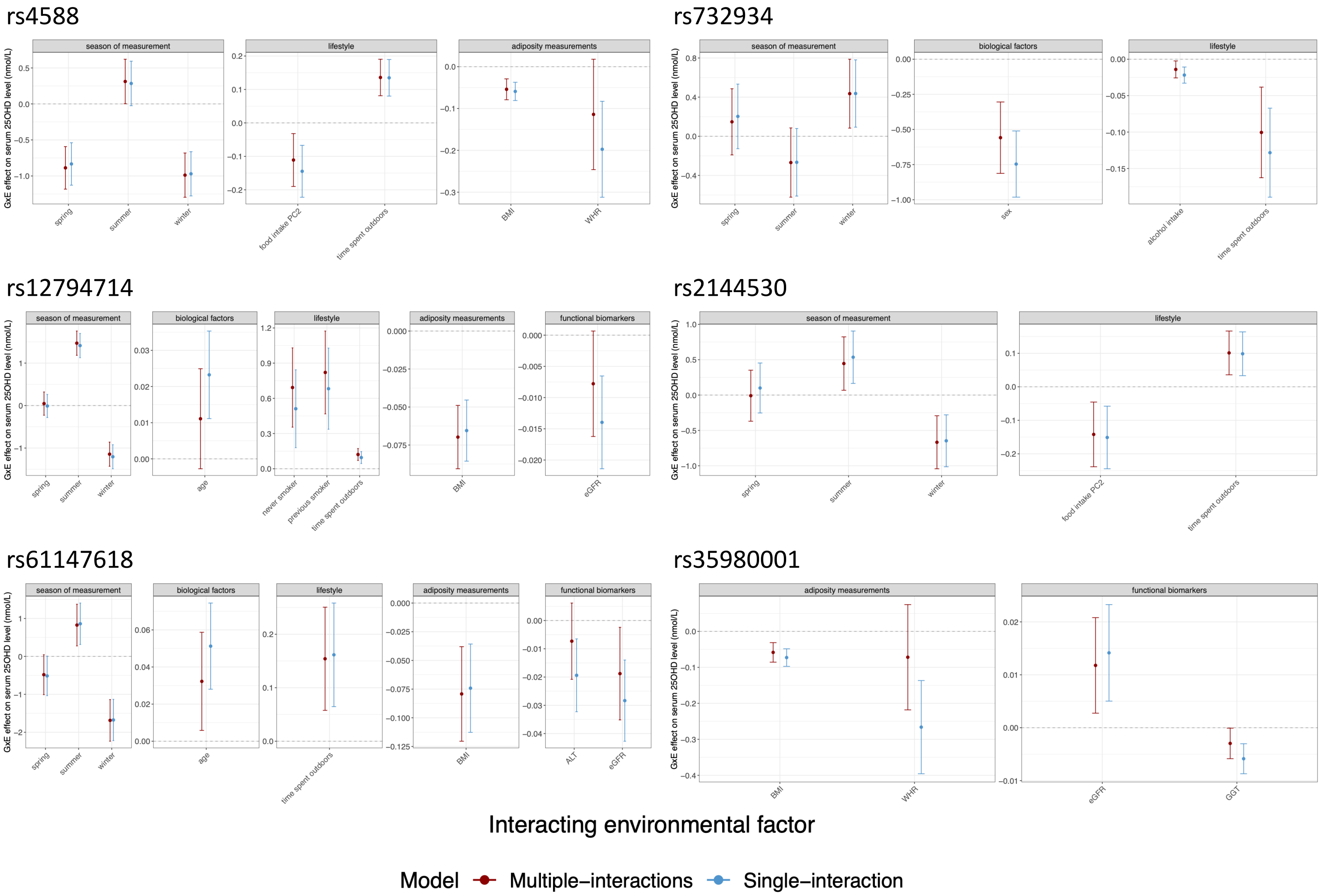


**Supplementary Figure S10.** Comparison of interaction effect estimates in multiple-interactions models and single-interaction models.

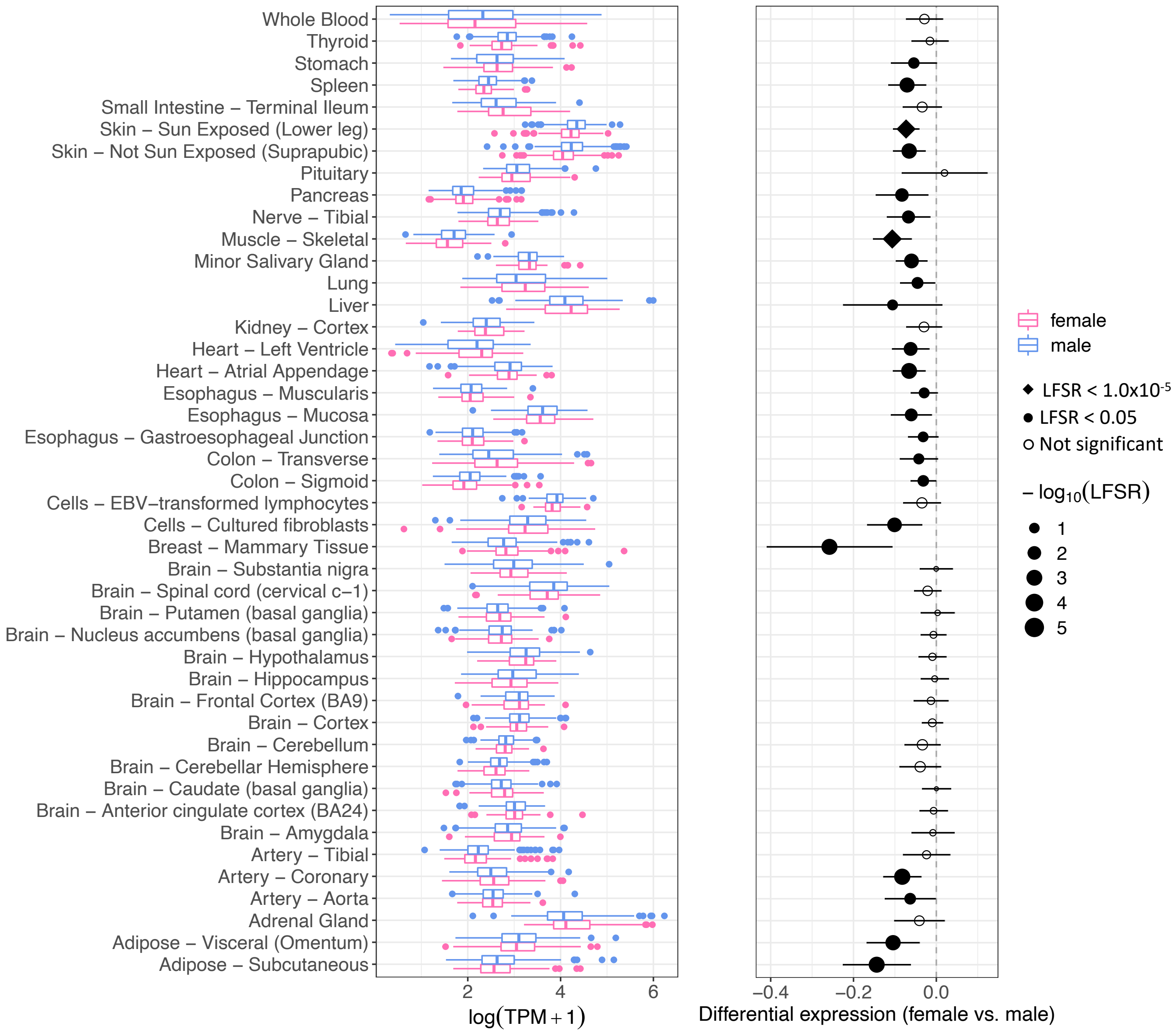


**Supplementary Figure S11.** Sex-differential expression of *DHCR7* detected in the GTEx. Normalized expression levels are illustrated for each GTEx tissue. Each box spans from the first quartile to the third quartile with the line inside the box representing the median and the whiskers representing 1.5x the inter-quartile range. In the illustration of differential expression, dots represent estimated effects of sex on gene expression. Error bars indicate 95% confidence intervals. A negative value denotes a higher average expression level in males. LFSR, local false sign rate; TPM, transcript per million.
